# Validation and Implementation of Real Time PCR for Rapid Identification of *Candida auris* from Clinical Surveillance Samples in a Local Public Health Laboratory Setting

**DOI:** 10.1101/2022.07.02.22277184

**Authors:** Lydia Mikhail, Megan Crumpler, Marie Bourgeois, Jill Roberts

## Abstract

**Background:** *Candida auris* is a worldwide emerging pathogen known for causing infections and outbreaks in health care settings. In Orange County, CA, *C. auris* has been circulating and causing outbreaks in long term care facilities since 2019, with a total of 1,017 cases detected from February 2019 to December 2021.

**Objective:** To evaluate an rt-PCR assay for rapid identification and assessment of colonization of *C. auris* from patient surveillance samples at the OC Public Health Laboratory.

**Method:** An extraction protocol using the MasterPure kit and automated EZ1 Advanced XL followed by rt-PCR using PerfeCTa Multiplex qPCR TaqMan, and the 7500 Fast Dx was conducted. The assay was evaluated using 131 previously confirmed patient samples and 123 prospective fresh samples from different body sites.

**Results:** The assay was highly reproducible at an LOD of 230 CFU/ml. The sensitivity and specificity of the 131 samples was 90 % and 86 % respectively and for the 123 samples it was 93% and 90% respectively. ROC analysis was calculated using all samples (n = 254) to determine the most ideal diagnostic value. The AUC was 0.922 and optimal cutoff was a cycle threshold 36.78. A CT ≤37.00 is the ideal diagnostic value for patient surveillance samples.

**Conclusion:** The successful implementation of the rt-PCR assay for *C. auris* in a local public health laboratory allows for accurate and rapid screening and identification of *C. auris* which can contribute to enhanced surveillance and control of *C. auris* spread in local health care facilities.

## Introduction

*Candida auris* is an emerging pathogenic yeast that has several unique features that have given it worldwide notoriety. It is inherently highly multidrug resistant (Forsberg et al., 2019), causes bloodstream (Chowdhary et al., 2013; Lee et al., 2011) and urine infections (Belkin et al., 2018), leads to invasive disease and is associated with high mortality rates (Armstrong et al., 2019; Ruiz-Gaitán et al., 2018). It causes serious nosocomial infections and outbreaks in patients that are critically ill or highly dependent, and among those staying in long-term health care facilities or the ICU (Calvo et al., 2016; Das et al., 2018; Sears & Schwartz, 2017).*C. auris* was first identified in 2009 from an ear swab of a patient in Japan (Satoh et al., 2009) and spread worldwide across over 20 countries spanning five continents from 2009 to present [Centers for Disease Control and Prevention (CDC), July 31, 2020)]. In Orange County, CA, *C. auris* has been circulating and causing outbreaks in long term care facilities since 2019, with a total of 1,017 cases detected from February 2019 to December 2021.

Outbreak investigations commonly start with laboratory testing (Adams et al., 2018). Rapid and timely laboratory diagnosis of infectious diseases are imperative for proper patient care and outcome, expedient public health intervention and effective infection control measures. However, emerging pathogens may be difficult to identify when they are first discovered as is the case with *C. auris*. Challenges are encountered in the laboratory identification of *C. auris* due to underreporting, misidentification (Lockhart, Berkow, Chow, & Welsh, 2017) and the requirement of specialized testing methods (Vallabhaneni et al., 2016).

The laboratory methods of identification available for *C. auris* include phenotypic or morphological characteristics in culture, biochemical based systems, mass-spectrometry (MALDI-TOF), polymerase chain reaction (PCR), DNA sequencing and whole genome sequencing (WGS).

Molecular techniques, such as PCR, are highly sensitive and specific and are more rapid, reliable and accurate than other available methods. The shorter turnaround time for *C. auris* identification in clinical and public health laboratories is of great advantage because this can help with preventing outbreaks and improving survival rates among patients by facilitating early treatment regimens (Kordalewska et al., 2017). Screening for *C. auris* using skin swabs is helpful in identifying asymptomatic colonized individuals and is often used for patients at high risk for acquiring *C. auris*, including residents at long-term care facilities. (Lockhart, Lyman, & Sexton, 2022).

The purpose of this study was to evaluate and validate real-time PCR (rt-PCR) based methods for screening of *C. auris* surveillance samples in a local public health laboratory setting to assess colonization. An additional goal was to apply findings to develop best practice testing workflow(s) to daily laboratory routine protocols for rapid response to *C. auris* outbreaks.

## Ethics

Applications for this project were submitted to both the University of South Florida IRB Committee and IRB Safety. The IRB committee reviewed the project and a “Not Human Subjects Research Determination” waiver was granted on 5/4/2021.

## Methods

1. *C. auris* PCR validation
1.1 Samples. The PCR assay was initially evaluated using 131 previously tested and frozen/refrigerated patient surveillance samples (60 negatives, 71 positives). These were obtained from the Washington State Public Health Laboratory with is designated by the CDC as an Antibiotic Resistance Laboratory Network laboratory (WA ARLN).
1.2 Prospective samples. Once validated, the rt-PCR assay was tested prospectively using 123 fresh surveillance samples collected from different body sites (nares, axilla, groin, perirectal, fingers/hands and axilla/groin) following the methodology of rt-PCR and culture previously described.
1.3 Specimens/samples - Patient swabs were collected in BD ESwab Collection and transport system with Amies buffer (BD Diagnostics, Sparks, MD). The swabs were vortexed, then 200µl of patient Amies buffer was transferred to a labelled aliquot tube.
1.4 DNA extraction. A novel semi-automated extraction protocol using the MasterPure Yeast DNA Purification kit (Biosearch Technologies, Hoddesdon, United Kingdom), and EZ1 Advanced XL (Qiagen, Hilden, Germany) instrument was performed. This combined purification and isolation method is referred to in this article as MPEZ1. The MasterPure Yeast DNA purification kit uses a non-enzymatic method for the lysis of fungi by a salting out procedure to precipitate proteins and an alcohol precipitation step to purify the DNA (Fredricks, Smith & Meier, 2005). For this *C. auris* extraction method, 200 µl of Amies was processed until the precipitation with MPC Protein Precipitation Reagent step found in the MasterPure Yeast DNA Purification Kit. The resulting supernatant was transferred to a clean EZ1 sample tube. The remainder of the extraction was performed on the EZ1 Advanced XL using the EZ1® DNA Tissue kit for automated purification of DNA.
1.5 *C. auris* real-time PCR assay. After extraction, real-time PCR was performed using PerfeCTa Multiplex qPCR TaqMan (Quanta Biosciences, Gaithersburg, MD), and the 7500 Fast Dx (Applied Biosystems, Waltham, MA). All primers and probes were obtained from Integrated DNA Technologies (idtdna.com) and were designed from the ITS2 gene region common to all four *C. auris* clades (Leach, Zhu, & Chaturvedi, 2018). Primers and probes were also acquired for an inhibition control from a Bicoid plasmid gene (Cat # 34340; Addgene, Watertown, MA; USA). Each sample was tested in a 96 well reaction plate with master mix containing 15 μl of 1x PerfeCTa Multiplex qPCR ToughMix (Quanta Biosciences), a 500 nM concentration of *C. auris* forward (V2424) and reverse (V2426) primers, and a 100 nM concentration of each *bicoid* forward (V2375) and reverse (V2376) primers, and a 100 nM concentration each of *C. auris* (V2425P) and *bicoid* (V2384P) probe (Leach, Zhu, & Chaturvedi, 2018). Each PCR run included 5 μl of positive extraction and amplification control (*C. auris* AR Bank #389; diluted at 10^−2^ −10^−3^); 5 μl of negative reagent control and 5 μl negative extraction control (200 µl of Amies buffer). Bicoid inhibition control plasmid DNA (1 μl per reaction) was added to the master mix after the negative reagent control was prepared.

The ABI 7500 Fast Dx (Applied Biosystems, Waltham, MA) cycling parameters were 95°C for 20 s, followed by 45 cycles of 95°C for 3 s and 60°C for 30 s. Specimens were reported as detected, not detected or indeterminate if PCR inhibition was observed.

1.6 Culture confirmation A 100 μl sample in Amies was inoculated to 2.0 mL Salt Sabouraud Dextrose (SSD) Broth (Welsh et al., 2017) for enrichment. The culture tubes were incubated by shaking at 200 rpm at 40°C and checked daily for turbidity/growth for up to 5 days. Once visible growth was detected (usually 48 hr), one CHROMagar® Candida (Becton Dickinson, Franklin Lakes, NJ) plate was inoculated. The CHROMagar media plates were incubated at 37°C for up to 2 days. If growth was indicative of *C. auris* (cream, pink or mauve colonies), identification was confirmed by MALDI-TOF Bruker Daltonics, Inc., Billerica, MA). For MALDI-TOF confirmation, yeast isolates were subcultured onto Sabouraud dextrose agar and incubated at 35 °C for 24–36 hours. Protein extraction was performed in a Biological Safety Cabinet using the CDC custom extraction method (https://www.cdc.gov/fungal/lab-professionals/yeast-maldi-tof.html) to optimize identification of yeast isolates that are represented in the Bruker Clinical Application and MicrobeNet databases (Frasier et al, 2015). Culture and rt-PCR results were compared to expected results.
2. Limit of detection (LOD). The LOD is the lowest concentration of *C. auris* repeatedly reproducible by the PCR screening assay. A panel was prepared using spiked samples with serial 10-fold dilutions of *C. auris* using *C. auris* CDC/AR bank #381 and # 389. From a 48-hour *C. auris* 381 culture isolate, 1-3 colonies were picked with a loop and suspended in a 3mL saline tube. A suspension was made and turbidity was measured using the Grantbio Den1B McFarland Densitometer (Grant Instruments, Cambridgeshire, UK) to equal 0.5 McFarland. Serial dilutions were made from 10^−1^ to 10^−5^, each dilution was plated to a SAB plate using a lawn streak, and incubated for 48 hours at 37°C. A volume of 200ul of each serial dilution sample was aliquoted to process for DNA extraction. All dilutions were tested in triplicate.
3. Analytical inclusivity and exclusivity. The *C. auris* rt-PCR assay analytical inclusivity and exclusivity validation panel consisted of 8 samples total, including 4 *C. auris* isolates representing the 4 clades (CDC/AR # 382, 381, 383, 385) and 4 non-*C*.*auris* (*C. duobushaemulonii, C. haemulonii, C. krusei* and *C. lusitaniae)*. Cell suspensions (within the optimal LOD range) of the isolates were prepared in ESwab Liquid Amies media.
4. Statistical analysis To assess the diagnostic value of the real-time PCR assay in *C. auris* detection from surveillance samples, the culture method was selected as the “gold standard.” A receiver operating characteristic (ROC) curve (SPSS version 28) was generated with the area under the curve (AUC) calculated for the rt-PCR results. The P values were set at confidence interval of 95%.

## Results

### Accuracy

WA ARLN provided 131 validation specimens (60 negatives, 71 positives) for use in this analysis. There was 91% agreement between the rtPCR results conducted at OCPHL and the WA positive and negative specimens (Table 1).

**Table 1:** Comparison of *C. auris* detection by rt PCR at OCPHL vs. WA ARLN

|  | WA positive | WA neg | Total | Accuracy |
| --- | --- | --- | --- | --- |
| OCPHL positive | 59 | 0 |  |  |
| OCPHL negative | 12 | 60 |  |  |
| Total | 71 | 60 | 131 | 91% |

### LOD

To determine the detection limits of the assay, 2 *C. auris* isolates (CDC/AR # 389 and 389) diluted from 10^−1^ to 10^−5^, were tested in triplicate.

The LOD was calculated based on CLSI MM17 Validation and Verification of Multiplex Nucleic Acid Assays (CLSI, 2007), where the LOD is estimated based on the number of positive replicates. The LOD results for *C. auris* # 381 = 190 CFU/mL (average of 80, 230, 260 CFU/mL). *C. auris* # 389 = 270 CFU/mL (average of 360, 320, 130 CFU/mL). The average LOD for this *C. auris* PCR assay = 230 CFU/mL Table 2 summarizes the LOD data for *C. auris* # 381 and #389 replicates at different dilutions.

**Table 2:** LOD data for *C. auris* # 381 and #389 replicates at different dilutions

|  | Dilutions |  |  |  |  |  |
| --- | --- | --- | --- | --- | --- | --- |
| *C. auris* | 1:1000000 | 1:100000 | 1:10000 | 1:1000 | 1:100 | 1:10 |
|  | % Replicate Testing Positive |  |  |  |  |  |
| 381 | 100 | 100 | 100 | 100 | 100 | 0 |
| 389 | 100 | 100 | 100 | 100 | 100 | 33 |

### Analytical inclusivity and exclusivity Validation

The *C. auris* rt-PCR assay analytical inclusivity and exclusivity validation panel consisted of 8 CDC/AR samples total, including 4 *C. auris* isolates (4 clades) and 4 non-*C*.*auris* (*C. duobushaemulonii, C. haemulonii, C. krusei and C. lusitaniae*).

All *C. auris* isolates were correctly identified by PCR, growth in SSD broth, on CHROMCandida agar and using MALDI-TOF. The results from the PCR assays were 100% concordant with culture results. The real-time PCR assay was highly specific, as none of the other organisms cross-reacted while *C. auris* organisms belonging to all known phylogenetic clades by whole-genome sequencing yielded positive results.

### Sensitivity and Specificity

Sensitivity and specificity entailed testing screening swabs collected from human patients using the *C. auris* rt-PCR assay. For the validation of the method, a total of 131 surveillance swabs were tested. The 131 validation samples were obtained from the WA ARLN consisting of 60 negatives and 71 positives. Samples were shipped on ice and the collection date was often > 96 hours. Samples were held at 4°C for an extended period.

The samples were tested by rt-PCR and inoculated to SSD broth. Positive growth on SSD broth was followed by culture on CHROMCandida agar and MALDI-TOF identification. Results were evaluated relative to the culture-based gold standard. The sensitivity and specificity were 90% and 86% respectively (Table 3)

**Table 3.** Comparison of *C. auris* detection by rt-PCR and culture (validation samples)

| Validation samples | *C. auris* detection by culture |  |  |  |  |
| --- | --- | --- | --- | --- | --- |
| *C. auris* detection by PCR | Positive | Negative | Total | Sensitivity | Specificity |
| Positive | 46 | 11 | 57 | 90 % | 86 % |
| Negative | 5 | 69 | 74 |  |  |
| Total | 51 | 80 | 131 |  |  |

Of the 131 samples tested, 51 were culture positive and of those 46 were also PCR positive. The remaining 80 were culture negative and 69 were also PCR negative. Regarding the 11 culture negative rt-PCR positive samples, a closer comparison was made to the original WA PCR results with OC results. 11/11 matched, giving an accuracy of 100%. This was sufficient evidence and acceptable for the low specificity of the test.

### Prospective samples

Once validated, the rt-PCR assay was performed for the 123 prospective fresh surveillance samples collected from different body sites (nares, axilla, groin, perirectal, fingers/hands and axilla/groin). The sensitivity and specificity of the 123 prospective samples were 93% and 90% respectively (Table 4).

**Table 4.** Comparison of *C. auris* detection by rt-PCR and culture (prospective samples)

| Prospective samples | *C. auris* detection by culture |  |  |  |  |
| --- | --- | --- | --- | --- | --- |
| *C. auris* detection by PCR | Positive | Negative | Total | Sensitivity | Specificity |
| Positive | 51 | 7 | 58 | 93% | 90% |
| Negative | 4 | 61 | 65 |  |  |
| Total | 55 | 68 | 123 |  |  |

### Statistical analysis

ROC curve analysis was used as a statistical tool for the diagnostic evaluation of the real-time rt-PCR assay A ROC curve was plotted by calculating the sensitivity and specificity of the rt-PCR cycle threshold (*CT)* values compared to culture results for swabs. Compared with the culture method, the areas under the ROC curves for the rt-PCR assay were tested twice and yielded 0.91 and 0.898 for the validation samples (n = 131). The corresponding *CT* values were 35.75 and 36.09 respectively. Therefore a *CT* of ≤36.00 was considered as positive and >36 as negative for this validation (Figure 1a).

**Figure 1a:**
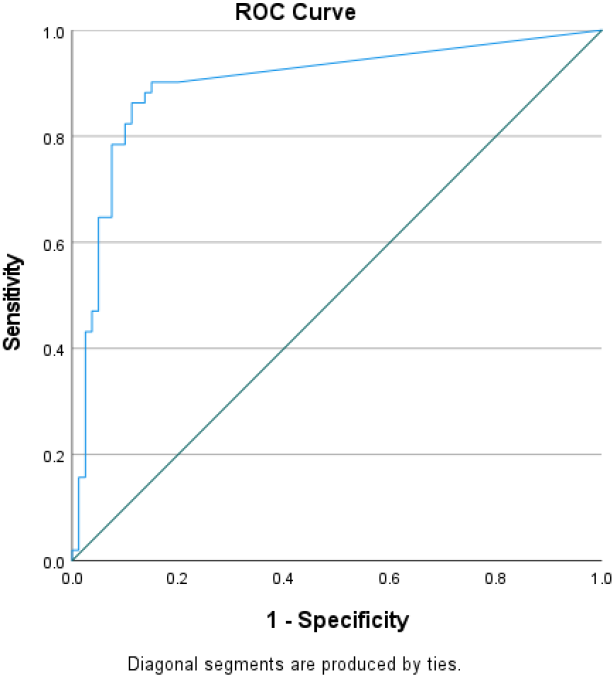
ROC curve analysis of *C. auris* real-time PCR assay for patient surveillance samples - Validation samples –AUC 0.89 and optimal cutoff 36.09

A revised ROC was performed for the 123 prospective samples yielding an AUC of 0.948 and optimal cutoff 36.92 (Figure 1b). Another ROC curve analysis was calculated using all samples (validation and prospective) for a larger sample volume (n = 254) and to determine the most ideal diagnostic value. The AUC was 0.922 and optimal cutoff was 36.78 (Figure 1c).

**Figure 1b.**
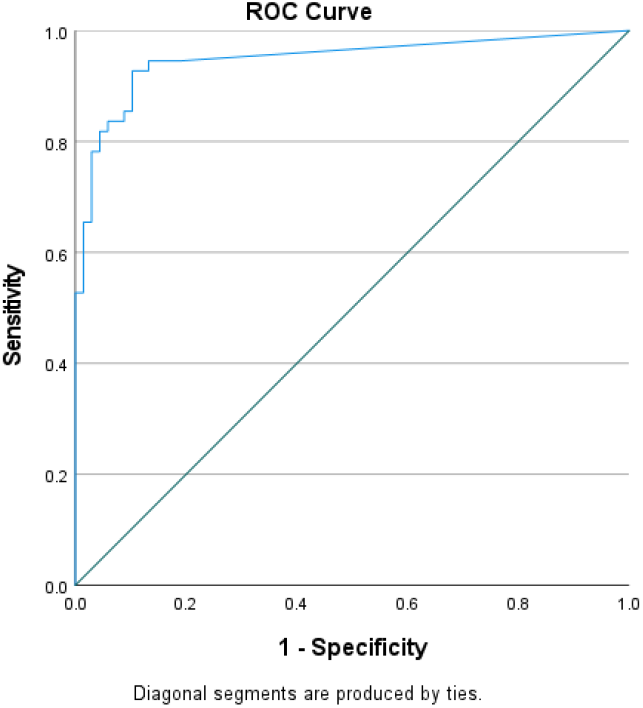
ROC curve analysis of *C. auris* real-time PCR assay for patient surveillance samples - Prospective samples – AUC 0.948 and optimal cutoff 36.92

**Figure 1c.**
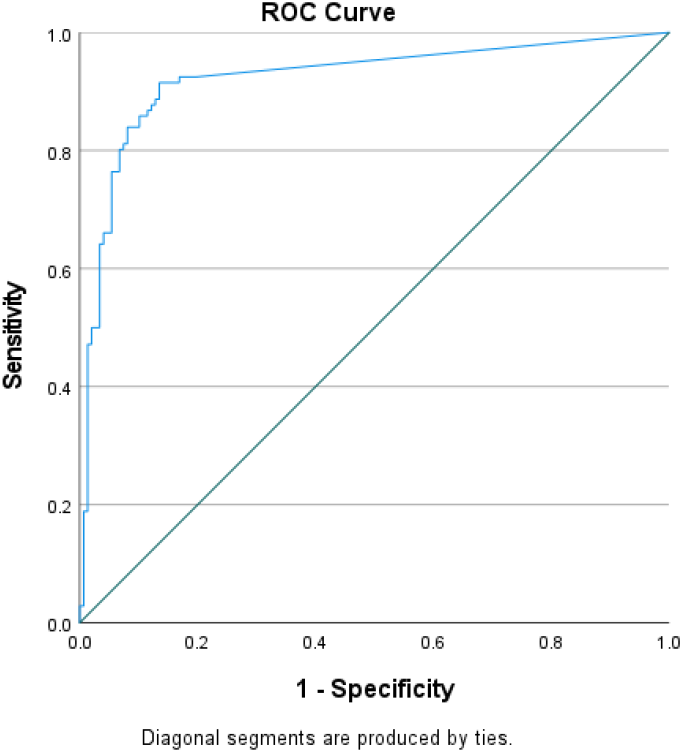
ROC curve analysis of *C. auris* real-time PCR assay for patient surveillance samples Pooled dataset – AUC 0.922 and optimal cutoff 36.78

Based on these results, we determined that a *CT* ≤37.00 is the ideal diagnostic value for patient samples. The cutoffs for reporting are detailed in Table 5.

**Table 5:**
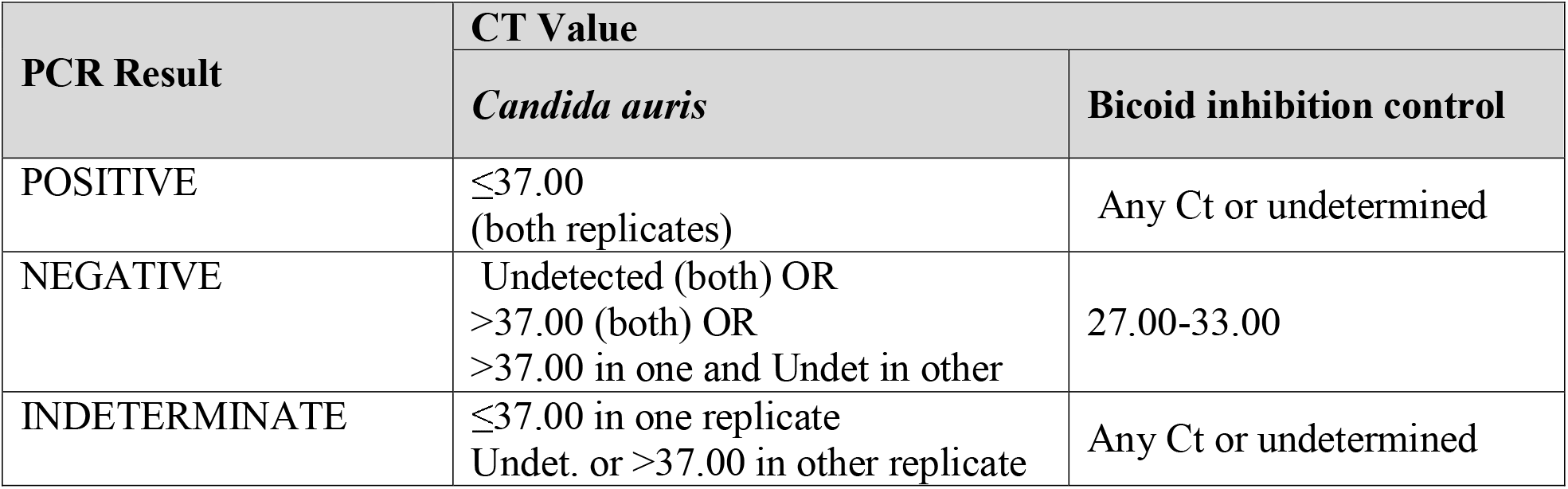
Interpretation of PCR Results

| PCR Result | CT Value |  |
| --- | --- | --- |
|  | *Candida auris* | Bicoid inhibition control |
| POSITIVE | $\leq 37.00$<br>(both replicates) | Any Ct or undetermined |
| NEGATIVE | Undetected (both) OR<br>>37.00 (both) OR<br>>37.00 in one and Undet in other | 27.00-33.00 |
| INDETERMINATE | $\leq 37.00$ in one replicate<br>Undet. or >37.00 in other replicate | Any Ct or undetermined |

### New workflow

Based on the results of the study, a new workflow was developed for testing surveillance samples from new admissions to SAUs and potential events/outbreaks (Figure 2).

**Figure 2:**
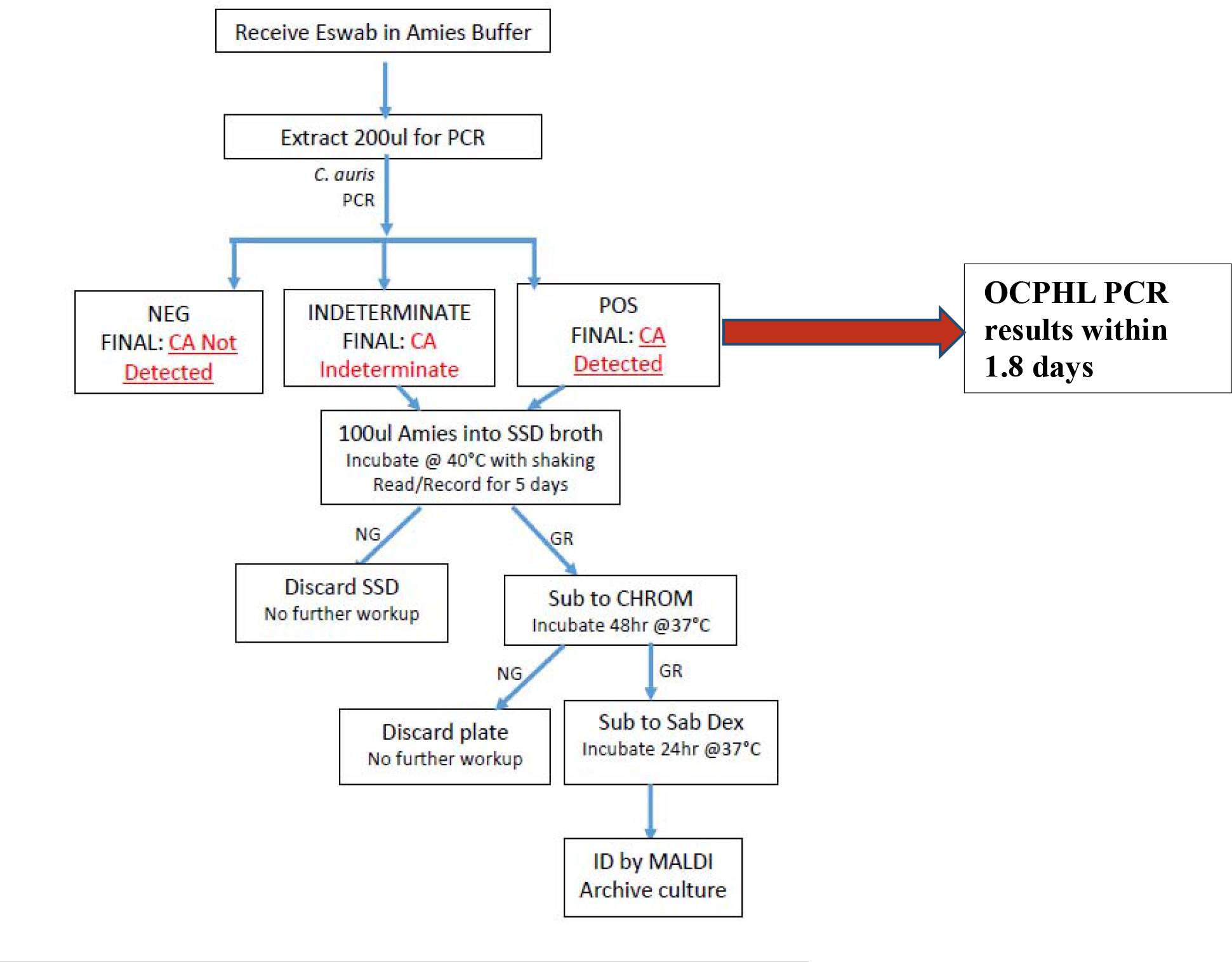
New workflow for *C. auris* surveillance swabs

OCPHL rt-PCR results were reported within 1.8 days on average compared to 8.3 days for surveillance testing at the ARLN lab. Culture confirmation was performed and isolates archived for future susceptibility and/or WGS.

## Discussion

The ongoing outbreak of *C. auris* that started in 2019 in local health care facilities in Orange County, CA, supported the need for laboratory methods for rapid and accurate means of diagnosis of this dangerous pathogen. This study describes a public health laboratory’s validation and implementation of rt-PCR to diagnose *C. auris* to enhance the OC Health Care Agency’s (OCHCA) infection control response and measures to prevent its spread.

### PCR advantages and limitations

The MPEZ1 is a novel semi-automated extraction method that takes 2 hours to extract nucleic acid from 12 samples and 2 controls. It is ideal for low sample volume because it averages 4 four hours from extraction to PCR This method also reduces errors associated with full manual processing using the MasterPure kit and also eliminates the issue of potential loss of the pellet that includes the nucleic acid, which could lead to false results.

Molecular techniques, such as rt-PCR are known to be fast, reliable and more sensitive and specific then other available methods. Rt-PCR is a powerful and accurate tool for laboratory identification of microorganisms because it is considered a culture-independent test (Kordalewska et al., 2017). For *C. auris*, swabs kept for a long time can be retrospectively screened for presence of *C. auris* by rt-PCR, but this can significantly reduce the viability of *C. auris* cells and chances for their recovery for culture (Kordalewska, & Perlin 2019).

This rt-PCR assay was validated using previously tested samples, prospective samples (Tables 3 and 4) as well as all four clades, for which all were correctly identified. For the four clades and non-*C*.*auris* isolates, there was 100% concordance of the PCR with culture/MALDI-TOF results. For the validation samples the sensitivity and specificity were 90% and 86% respectively (Table 3). For the prospective samples, the samples were 90% and 93% (Table 4). These results are somewhat lower than other PCR assays. For example in one PCR study conducted on 103 clinical isolates from axilla and groin skin swab samples, the qPCR technology used gave 93% sensitivity and 96% specificity (Sexton et al., 2018). Another study by Arastehfar et. al, (2019) demonstrated that the sensitivity and specificity for their PCR assay was 100% for both.

PCR can also detect DNA from both live and dead cells. For this evaluation, since culture is the gold standard, some of the rt-PCR results are considered either false positive or false negative. In both these cases, this could be due to unequal input volumes used for each method; 200 μl for PCR vs 100 μl for culture. An additional reason for false negatives could be due to incorrect holding or storage conditions. The less than ideal holding conditions were applicable to the validation panel samples as they were not tested within the 4-day recommended holding time.

The performance of this rt-PCT assay was deemed acceptable for rapid identification of *C. auris* and is ideal for local public health laboratory settings that are running small volumes and wanting quick turnaround time to results. In addition, this rt-PCR method can be applied to all sample types including the clinical, surveillance, and environmental samples.

### Benefit of rapid PCR screening workflow to OCHCA

According to the CDC, California is one of the top 4 states reporting > 101 clinical cases of *C*.*auris* from January 1, 2021 to December 31, 2021 (CDC, February 23, 2022). In Orange County, *C. auris* spread has been identified in several Long Term Acute Care Hospitals (LTACHs) and Ventilator-capable Skilled Nursing Facilities (vSNFs) since 2019. OCHCA recommends that facilities conduct screening of patients transferring from any Southern California LTACH or subacute units (SAU), and place them on appropriate empiric contact precautions while awaiting results. OCHCA continues to work with OC LTACHs and SAUs to conduct point prevalence surveys of *C. auris* on an ongoing basis and also responds to known exposure events.. A negative screening result can remove a patient from empiric precautions, but it may take several days to get the result (OCHCA, October 13, 2021).

To address this challenge, after the implementation of the rt-PCR, a new workflow was developed for testing surveillance samples from new admissions to SAUs and potential events/outbreaks (Figure 2). Reporting of results (detected, not detected or indeterminate) is made within 1.8 days on average compared to 8.3 days for surveillance testing at the regional WA ARLN. Culture set up and MALDI-TOF confirmation is still performed primarily to save isolates for future susceptibility testing and/or WGS.

Starting October 2021, additional surveillance samples were tested from new patient admissions and potential events/outbreaks in SAUs. Using the protocol described herein, a total of 310 new patient admissions and potential events/outbreaks in SAUs were tested from October 2021 to March 2022, with 14 positives, 2 indeterminate and a 1.8 day average TAT for PCR results. This successful implementation of rt-PCR for *C. auris* in the OCPHL accomplishes the goal of rapid identification of *C. auris* in individuals at long term health care facilities. This is advantageous for OCHCA to provide early detection, support containment of spread and institution of infection control measures and recommendations.

## Data Availability

All data produced in the present work are contained in the manuscript

